# Testing-isolation interventions will likely be insufficient to contain future novel disease outbreaks

**DOI:** 10.1101/2023.11.16.23298614

**Authors:** Jeffery Demers, William F. Fagan, Sriya Potluri, Justin M. Calabrese

## Abstract

When novel human diseases emerge into naive populations, identification and isolation of infected individuals forms the first line of defense against the invading pathogens^1,2^. Diagnostic testing plays a critical role^3,4^, but health agencies unprepared for a novel disease invasion may struggle to meet the massive testing capacities demanded by an epidemic outbreak^5^, potentially resulting in a failure of epidemic containment as with COVID-19^6^. What factors make a disease controllable versus uncontrollable with limited testing supplies remains unclear. Specifically, is the failure of testing-isolation unique to COVID-19, or is this a likely outcome across the spectrum of disease traits that may constitute future epidemics? Here, using a generalized mathematical disease model parameterized for each of seven different human diseases, we show that testing-isolation strategies will typically fail to contain epidemic outbreaks at practicably achievable testing capacities. From this analysis, we identify three key disease characteristics that govern controllability under resource constraints; the basic reproduction number, mean latent period, and non-symptomatic transmission index. Interactions among these characteristics play prominent roles in both explaining controllability differences among diseases and enhancing the efficacy of testing-isolation in combination with transmission-reduction measures. This study provides broad guidelines for managing controllability expectations during future novel disease invasions, describing which classes of diseases are most amenable to testing-isolation strategies alone and which will necessitate additional transmission-reduction measures like social distancing.

---

Invasions of novel diseases into naive human populations have had devastating consequences throughout human history, exemplified most recently by the COVID-19 pandemic^7^. Given the increasing potential for zoonotic spillover events driven by human activity^8,9^, novel pathogen emergence remains an ongoing threat that will continue to demand public health interventions. Identification and isolation of infected individuals plays a central role in responses to emerging epidemics^1,2^, acting to break transmission chains and limit population-wide disease spread. This control strategy utilizes diagnostic testing to identify infected cases^3,4^, most notably for diseases presenting nebulous or non-existent symptoms like COVID-19. Unlike lengthy development times required for vaccinations^10^, diagnostic tests can be developed rapidly after a pathogen has been genetically sequenced and subsequently deployed during the critical initial stages of an emerging epidemic^11^. The prominence of testing-isolation in public health management is underscored by the World Health Organization’s key message of “Test, Test, Test,” during the initial months of the COVID-19 pandemic^3^.

Despite the potential for rapid development, timely mass deployment of diagnostic tests early in an epidemic presents considerable logistical challenges. Not only must industrial power be leveraged to rapidly accelerate test production, the testing resources thus produced must be efficiently distributed and health care infrastructure expanded to accommodate large-scale test administration and specimen processing^12^. During the initial phases of the COVID-19 outbreak, testing capacity fell well short of demand, thereby resulting in severe bottlenecks to both test access and test processing times worldwide^5,13^. Testing backlogs were so severe that many regions were forced to institute strict resource rationing policies prioritizing front-line workers, vulnerable individuals, and severely ill patients^14,15^. These testing shortfalls drastically limited the rates at which infected individuals could be identified and consequently undermined the efficacy of identification-isolation policies^6,16^. Ultimately, identification-isolation strategies proved inadequate to stop the worldwide spread of COVID-19, with the disease eventually infecting over 700,000,000 people and claiming nearly 7,000,000 lives^17^.

The failure of testing-isolation to contain COVID-19 raises urgent questions for public health management. Was the inefficacy of resource-constrained testing-isolation unique to the particular properties of COVID-19, or is this a typical outcome to be expected for a variety of diseases? More generally, what characteristics make a disease controllable or uncontrollable under resource limitations, and how do these factors influence the resource levels required to thwart epidemic invasions? Most importantly, what if any classes of diseases are actually controllable at practicably achievable testing capacities? Given that the logistical barriers to testing capacity build-up are not unique to COVID-19, resource shortfalls can be anticipated for future outbreaks of novel diseases. Answers to these questions are therefore critical for both planning epidemic responses and informing expectations on what can realistically be achieved given real-world testing limitations.

Previous theoretical investigations have analyzed controllability over landscapes of disease characteristics for non-pharmaceutical interventions like contact tracing with quarantine and symptom-based self-isolation^18,19^. Principal findings indicate that a disease’s natural history, inherent transmissibility, and degree of non-symptomatic transmission are key determinants of intervention efficacy. However, these pre-2019 studies could not have anticipated the unprecedented inadequacies in health-care infrastructure that limited the efficacy of non-pharmaceutical interventions during the initial COVID-19 outbreak. As such, these pre-pandemic findings did not provide direct insight into the feasibility of managing COVID-19 based on relevant practical quantities like testing capacity and were ultimately unable to guide critical decisions for public health measures. More generally, it is not known how generic disease controllability assessments from previous studies translate to actionable policy advice for the resource-constrained testing-isolation scenarios likely to occur during future novel disease outbreaks. To address these critical knowledge gaps, we analyze controllability for seven focal diseases exhibiting a wide array of characteristics and natural histories using a generalized mathematical model for infectious disease spread that incorporates explicit testing and isolation interventions, resource constraints, and optimizable resource management strategies. We determine the testing capacities required to contain epidemic invasions for each pathogen and identify the key characteristics that govern controllability across diseases under resource constraints. We find that interactions among three key disease characteristics (the basic reproduction number, mean latent period, and non-symptomatic transmission index, all defined below) strongly influence disease controllability and explain differences in intervention outcomes for COVID-19 versus previous diseases. Our results provide realistic controllability expectations for future novel pathogen invasions that can guide decision-making for more invasive non-pharmaceutical interventions like lockdowns.

## Results

### Controllability across a spectrum of diseases

Ranges of epidemiological parameter values for the original, Delta, and Omicron COVID-19 variants along with the diseases smallpox, measles, SARS, and hepatitis A (excluding the influence of vaccines where possible) were selected through literature searches of published data and mathematical modeling studies (Methods, Extended Data Table 1, Extended Data Table 2). From these ranges, 6000 parameter sets for each disease were drawn using Latin hypercube sampling. Parameter sets were then input into a generalized mathematical model for infectious disease spread. Key model elements include testing and isolation control strategies, tunable testing efficiency from high-efficacy contact-tracing to low-efficacy randomized sampling, limited testing capacity, and optimizable resource management decisions^20^ (Supplementary Information). Model output was used to calculate the minimum testing capacity required to prevent disease invasions into naive populations as determined by the controlled disease reproduction number^21^ (Methods). The calculated minimal testing capacities define thresholds for disease controllability; testing and isolation measures have sufficient resources to halt the initial exponential growth of an epidemic only if testing capacity meets or exceeds the threshold capacity. Threshold testing capacities also provide simple measures of relative controllability across diseases; the larger the threshold, the more difficult a disease is to control.

Our overall results on controllability are generally consistent with real-world experience for disease management outcomes (Fig. 1a). For example, the median threshold testing capacity for the original COVID-19 strain suggests that at least 30 tests per thousand per day would have been required to contain the initial epidemic (Fig. 1a). This result is consistent with the observed rapid initial spread of COVID-19 despite the early implementation of test-trace-isolation programs, given that most countries were limited by testing capacities below 5 tests per thousand per day^17^. Consequently, absent ubiquitous compliance with lockdowns, social distancing, or other restrictive measures, the worldwide spread of COVID-19 was all but inevitable under the resource constraints of the early pandemic. The Delta and Omicron COVID-19 variants are even less controllable than the original strain by a factor of 3 to 9 (Delta and Omicron median controllability thresholds, Fig. 1a), consistent with their rapid spread rates despite greater test availability when those variants emerged^17^. The Delta variant in particular exhibits a level of controllability commensurate with that of measles (Fig. 1a), suggesting that the inability to prevent the rapid spread of Delta COVID-19 is comparably difficult to suppressing measles outbreaks during the pre-vaccine era. Smallpox and hepatitis A are more controllable than the original COVID-19 strain, but still difficult to control as they would have required approximately 10 tests per thousand per day or more to suppress in the pre-vaccine era (Fig. 1a). The 2003 SARS coronavirus, however, is exceptionally controllable compared to the other diseases (Fig. 1a), which is consistent with the relative ease with which the 2002-2003 epidemic was contained.

**Fig. 1:**
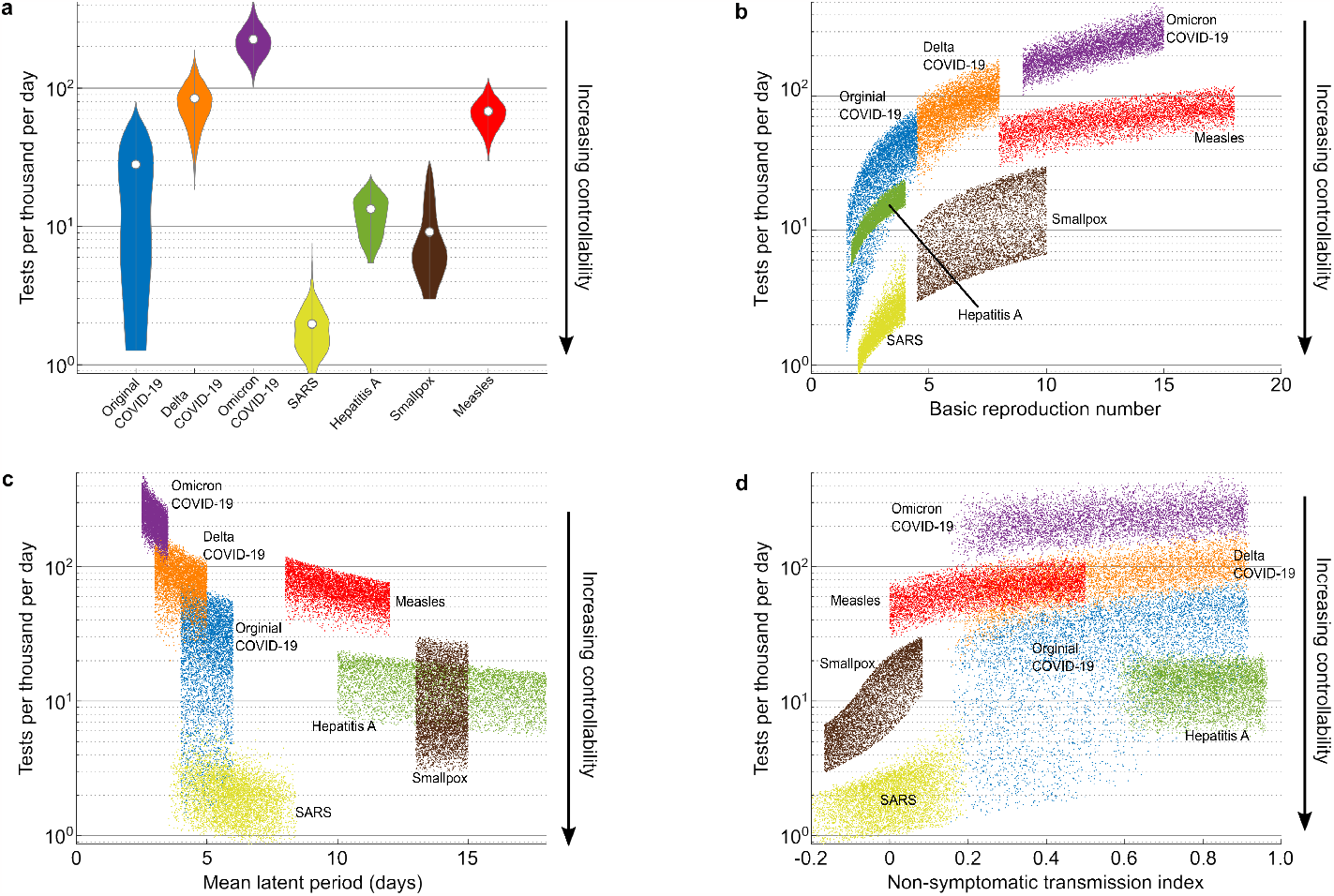
Controllability of seven past and current diseases quantified by threshold testing capacities. Log-scaled y-axes in all plots indicate threshold testing capacities for suppressing or preventing disease outbreaks in units of tests administered and processed per thousand individuals per day. Smaller thresholds indicate greater degrees of controllability because fewer testing resources are required to prevent large scale disease invasions. **a**, Violin plots of controllability thresholds for 6000 points sampled from the parameter spaces of each disease, where white dots indicate median controllability levels. **b – d**, Controllability thresholds plotted as a function of key disease characteristics (basic reproduction number, mean latent period, non-symptomatic transmission index) for each disease, where each point represents a particular combination of parameters sampled from a disease’s parameter space.

### Key determinants of disease controllability

Three disease characteristics primarily determine controllability: the basic reproduction number *R*_0_, the mean latent period *τ*_*L*_, and the non-symptomatic transmission index NSTI (Figs. 1b–1d). The basic reproduction number *R*_0_ (Methods) represents the average number of secondary infections generated by an individual over the course of their infection in an otherwise susceptible population without any intervention measures^21^. This factor is a proxy measure for overall transmission risk, where larger *R*_0_ values generally indicate diseases that are expected to be more contagious and infect more people^22^. As expected, diseases with larger *R*_0_ tend to be less controllable than diseases with smaller *R*_0_ (Fig. 1b). However, the original COVID-19 strain, SARS, and hepatitis A all share comparable *R*_0_ ranges, yet they exhibit substantial differences in medians and ranges of controllability (Figs. 1a, 1b). Likewise, the Delta COVID-19 variant and smallpox share comparable *R*_0_ ranges yet exhibit substantial differences in controllability. The *R*_0_ range for measles extends beyond that of the Omicron COVID-19 variant, yet Omicron is significantly less controllable than measles. These results indicate that although *R*_0_ is an important factor, it is not enough to determine disease controllability.

The mean latent period *τ*_*L*_ (Methods) describes the expected time elapsed between the initial infection of an individual and their onset of infectiousness. This period provides a window of time to identify and isolate new cases before they have an opportunity to generate new transmissions. Intuitively, a disease that leaves less time to identify and isolate an infected individual before they begin infecting others will be more difficult to contain, so shorter latent periods tend to reduce the controllability of a disease (Fig. 1c). However, measles and hepatitis A share a range of *τ*_*L*_ values over which they exhibit significant differences in controllability, as do the COVID-19 variants (Fig. 1c). Moreover, the original COVID-19 strain and SARS exhibit significant differences in controllability over their common *τ*_*L*_ range (Fig. 1c) despite their comparable *R*_0_ values (Fig. 1b). Thus, neither *R*_0_ nor *τ*_*L*_, alone or in conjunction with one another, are sufficient to determine disease controllability.

NSTI (Methods) measures the degree to which non-symptomatic transmission occurs for a disease (whether from pre-symptomatic cases or cases that remain asymptomatic throughout the course of an infection). Larger positive values indicate diseases for which larger fractions of new infections are generated by pre- or asymptomatic cases, while negative values indicate diseases for which symptom onset occurs well before the onset of significant levels of contagiousness and for which few or no asymptomatic cases occur. As NSTI increases towards one, the infected population becomes more difficult to distinguish from the general population, so larger NSTI values are expected to reduce controllability (Fig. 1d). However, although all three COVID-19 variants share similar NSTI levels and some overlap in *τ*_*L*_ values, they exhibit vastly different levels of controllability. Moreover, measles and the Omicron variant share a range of similar NSTI values, with measles having comparable or larger *R*_0_ values, yet Omicron is less controllable than measles. Hepatitis A has a larger NSTI than measles, yet measles is less controllable than hepatitis over their common range of *τ*_*L*_ values. Thus, NSTI alone or in conjunction with *τ*_*L*_ or with *R*_0_ is insufficient to explain disease controllability.

Only by considering *R*_0_, *τ*_*L*_, and NSTI together can the relative controllability of diseases be explained. Figure 2 plots the median values of these characteristics for each disease. Each plot represents a combination of two of the three characteristics; the quadrants of characteristic pair values that most severely reduce controllability are highlighted by red boxes. The Omicron COVID-19 variant is the only disease present in these worst-case quadrants for all characteristic pairs, thus corresponding with it being far less controllable than all other diseases in our study. Further, we find that the significantly larger controllability of SARS compared to the original COVID-19 strain over their common *τ*_*L*_ range is due to COVID’s much larger NSTI values, as the two diseases have comparable *R*_0_ ranges. Likewise, the greater controllability of hepatitis A compared to the original COVID variant is primarily driven by hepatitis A’s larger *τ*_*L*_, as both diseases share comparable NSTI and *R*_0_ ranges (although the marginally larger NSTI of hepatitis A may play a minor role as well). Further, the greater controllability of measles compared to the Omicron COVID variant is primarily driven by the former’s larger *τ*_*L*_, with Omicron’s marginally larger NSTI also playing a minor role.

**Fig. 2:**
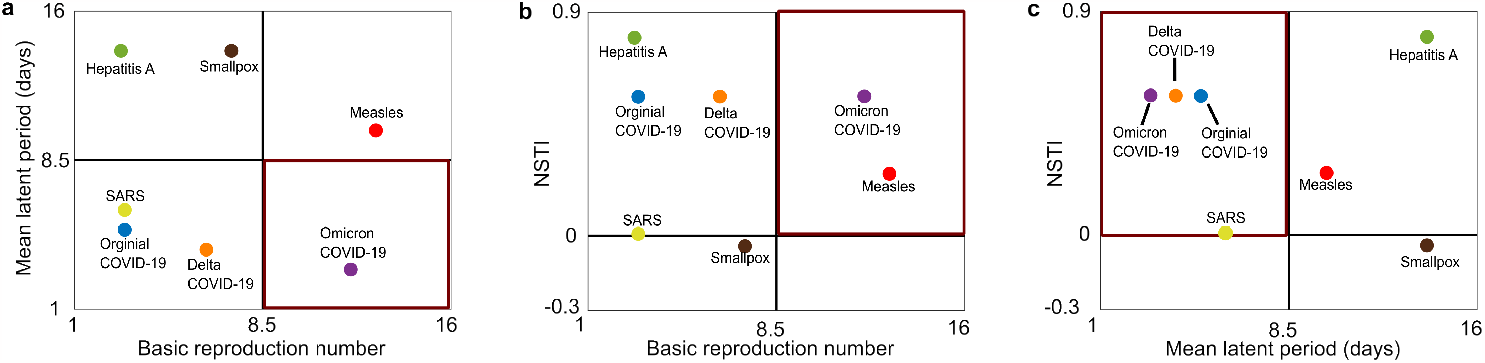
Median values of key disease characteristics (basic reproduction number, mean latent period length, and non-symptomatic transmission index). Values are projected onto two-dimensional sub-spaces of the three-dimensional disease characteristic space. Quadrants outlined in red represent the most difficult regions of 2D characteristic space to control, comprising large basic reproduction numbers, short latent periods, and positive NSTI.

### Compounding influences of interacting disease characteristics

Beyond their individual influences, interactions among *R*_0_, *τ*_*L*_, and NSTI are also important in explaining disease controllability. These interactions appear as differences in marginal controllability costs (Fig. 1), i.e. the rate of threshold testing capacity increase (or controllability decrease) in response to changes in one of the key disease characteristics. For example, the marginal controllability cost of increasing *R*_0_ is greater for the Omicron COVID-19 variant than for measles, as evidenced by Omicron’s larger slope of threshold testing capacity plotted against *R*_0_ (Fig. 1b). This difference is due to Omicron’s much smaller *τ*_*L*_ and marginally larger NSTI (Figs. 1c and 1d). These factors exacerbate the degree to which larger *R*_0_ values become more costly to control, thus indicating possible *R*_0_ : *τ*_*L*_ and *R*_0_ : NSTI two-way interactions and a possible three-way interaction among all key disease characteristics. A *τ*_*L*_ : NSTI interaction is evident when comparing SARS to the original COVID-19 strain. Here, COVID’s greater marginal controllability cost of *τ*_*L*_ reduction within their common *τ*_*L*_ range (Fig. 1c) is attributed to its larger NSTI (Figs. 1d), suggesting that large NSTI values can exacerbate the degree to which a short *τ*_*L*_ reduces controllability. Overall, these results suggest that interactions among *τ*_*L*_, NSTI, and *R*_0_ compound their influences on disease controllability, where combinations of problematic values of multiple key disease characteristics (i.e., the red regions in Fig. 2 or combinations thereof) can reduce controllability to a greater degree than would be expected from their combined individual influences.

The seven diseases we consider differ qualitatively by a number of factors aside from the key disease characteristics. For example, the Omicron variant incorporates an asymptomatic class while measles does not (Extended Data Table 1). To verify that the observed differences in marginal controllability cost are indeed attributed to interactions between the key disease characteristics rather than differences among other factors, we consider a single generically parameterized disease with wide parameter ranges representative of the real diseases in our study (Extended Data Table 1). We again utilize Latin hypercube sampling to calculate controllability threshold testing capacities as a function of the key disease characteristics (Extended Data Figs. 1a-1c). We then use multiple linear regression analysis to fit threshold testing capacities to the independent variables *R*_0_, *τ*_*L*_, and NSTI, considering a nested set of regression models with no interactions, two-way interactions, and both two and three-way interactions (Methods).

Two and three-way interactions contribute significantly to regression model performance (F-statistic nested model comparison), while model selection criteria (AIC and BIC) overwhelmingly select for the full model with all interactions (Extended Data Table 3). With all interactions included, the three key disease characteristics explain over 90% of the observed variance in threshold testing capacities (R-squared, Extended Data Table 3). In the full regression model, marginal costs provide direct insight into the influence of interactions on disease controllability (Methods). We find, for example, that the marginal controllability cost of latent period reduction increases with *R*_0_ and NSTI due to the *R*_0_ : *τ*_*L*_ and *τ*_*L*_ : NSTI interactions, respectively, and does so at enhanced rates for simultaneously large NSTI and *R*_0_ due to the three-way interaction (Fig. 3a). Analogous results follow for the marginal cost of NSTI increase relative to the *R*_0_ : NSTI and *τ*_*L*_ : NSTI interactions, with enhanced rates of growth for simultaneously small *τ*_*L*_ and large *R*_0_ (Fig. 3b). These results show directly how controllability losses due to short *τ*_*L*_ and large NSTI are worse for large *R*_0_, and how these effects compound when all three characteristics take on problematic values (large *R*_0_ and NSTI, small *τ*_*L*_). Conversely, when *R*_0_ is reduced, not only does a small *τ*_*L*_ itself become less problematic (smaller marginal costs), the value of NSTI becomes inconsequential to the degree of controllability loss associated with *τ*_*L*_ as indicated by convergence of the full interaction regression lines (Fig. 3a). Likewise, reducing *R*_0_ reduces both the impact of large NSTI and diminishes the influence of *τ*_*L*_ on the associated controllability loss (Fig. 3b). In effect, small *R*_0_ values can counteract the influence of the *τ*_*L*_ : NSTI two-way interaction by way of the three-way interaction. We thus find that in addition to reducing overall transmission risk, reducing *R*_0_ carries the added benefit of mitigating the effects of small *τ*_*L*_ and large NSTI as well as their compound influences. Correspondingly, explicit *R*_0_ reductions can markedly enhance controllability of the seven focal diseases. For example, explicitly reducing the sampled *R*_0_ values by a factor of 1/3 lowers the median controllability thresholds of hepatitis A and the original COVID-19 strain below 10 tests per thousand per day and below 5 tests per thousand per day for smallpox (Extended Data Fig. 2a). Analogous results hold for the compounding / mitigating influences of one disease characteristic on the others for all permutations of *R*_0_, *τ*_*L*_, and NSTI (Extended Data Fig. 1d-1i)

**Fig. 3:**
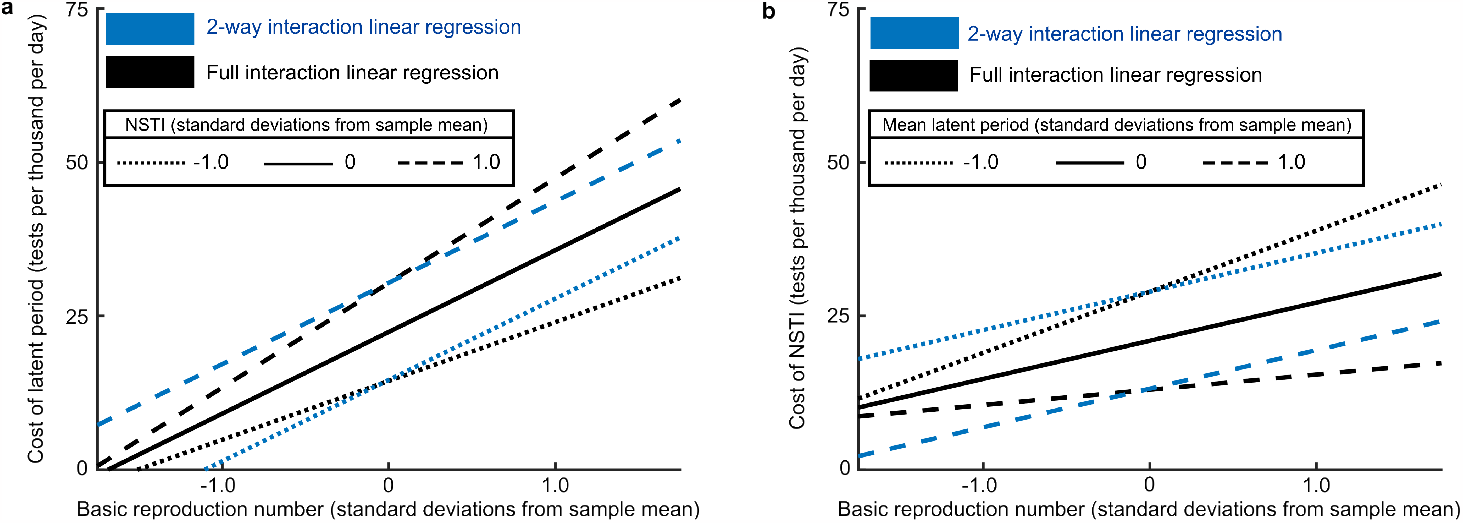
Marginal controllability costs for a generically parameterized disease. Standardized units are adopted for the basic reproduction number (mean 8.25, SD 3.90), mean latent period (mean 6.00 days, SD 1.73 days), and NSTI (mean 0.51, SD 0.23). Blue lines indicate marginal controllability cost calculated from the two-way interaction regression model, while black lines indicate the full two and three-way interaction model. Differences between corresponding blue and black lines indicate the influence of the three-way interaction term. **a**, Marginal controllability cost of latent period reduction for different values of *R*_0_ (x-axis values) and NSTI (separate lines). **b**, Marginal controllability cost of NSTI increase for different values of *R*_0_ (x-axis values) and *τ*_*L*_ (separate lines).

## Discussion

For a diverse set of disease traits exemplified by seven major human pathogens, resource-constrained testing-isolation interventions will generally struggle to prevent, suppress, or otherwise control an epidemic invasion. In particular, the maximum testing rates achieved by most countries over the first wave of the COVID-19 pandemic^17^ fall well below testing capacity thresholds required to halt disease invasion for all studied pathogens except SARS. Thus, the hope that testing-isolation could offer sufficient protection against COVID-19 spread by autumn 2020 to safely re-open schools and businesses without strict social distancing and behavior restrictions^12,23^ was never realistic. The inability to prevent outbreaks with limited testing supplies is not unique to COVID-19, but rather, is the typical situation across a broad spectrum of human pathogens. For some diseases like measles, epidemic containment requires enormous testing rates that may be unachievable even in highly developed countries. Moreover, our results represent optimistic estimates for controllability that assume efficacious wide-reaching contact tracing programs, perfect test sensitivity, perfect test specificity, perfect isolation for test-positive individuals, possible disease detection throughout the entire course of an infection, and optimal resource management decisions. Such ideal conditions are never achievable in the real world. In more realistic situations, prospects for controllability worsen considerably (a pessimistic scenario assuming low-efficacy random sample testing rather than contact tracing generally exhibits lower controllability across all diseases, but does not otherwise alter our principal results and conclusions (Extended Data Figs. 2b, 3, 4)). This study shows definitively that at the low levels of testing capacity expected during the initial phases of a novel disease outbreak, epidemic containment with testing-isolation alone is an exceptional rather than expected outcome.

Three key disease characteristics explain controllability trends under testing-isolation strategies: the basic reproduction number *R*_0_, the mean latent period *τ*_*L*_, and the non-symptomatic transmission index NSTI. These quantities contribute to the success or failure of non-pharmaceutical interventions like contact tracing-based quarantine and symptom-based self-isolation^18,19^, but this study makes clear exactly how these measures, and in particular the interactions among them, determine the initial spread or decay of a novel disease under resource constraints. Our study uniquely phrases these results directly in terms of a real-world measure for the capability of public health initiatives, i.e., testing capacity. This novel feature provides a crucial bridge between theoretical assessments of controllability over disease landscapes and the practical action-oriented priorities of public policy makers. From this study, public health agencies can gauge the feasibility of containing an emerging epidemic given available testing capacities once estimates for the three key disease characteristics are available. Correspondingly, our study sets a clear agenda for prioritizing epidemiological research during novel disease outbreaks. Estimated parameter ranges for *R*_0_, *τ*_*L*_, and NSTI should be obtained as soon as possible to determine prospects for epidemic containment and to set target goals for increasing testing capacity. Early COVID-19 research focused heavily on determining *R*_0_ values^24,25^ and the contributions of non-symptomatic transmission to disease spread^26,27^ (among other quantities), but to the best of our knowledge, estimates for the actual distribution of latent period values were only first published over a year into the pandemic^28^. Improved epidemic responses may result if latent periods receive increased attention early in an outbreak.

The prominence of the key disease characteristics in explaining controllability occurs because delays in the identification of infected individuals are a direct result of shortfalls in testing capacity. Specifically, individuals must wait longer to obtain tests and receive results when fewer testing resources are available, thus implying that lower testing capacities will cause longer delays. The three key disease characteristics determine the degree to which these delays will undermine disease containment efforts. Specifically, a decrease in *τ*_*L*_ will increase the window of time over which supply-related identification delays permit pre-isolation disease transmission. Likewise, an increase in NSTI implies that greater numbers of non-symptomatic (and thus non-infected) individuals must be tested to find sufficiently large numbers of active cases. This leaves fewer resources available for those who are actually infected, which consequently lengthens supply-related identification delays and increases the window of time for pre-isolation disease transmission. In either case, a larger risk of pre-isolation disease transmission reduces the overall level of epidemic control attainable with a given testing capacity. An increase in *R*_0_ implies that delays or failures in identifying infected cases will result in more secondary transmissions and thus further diminish the attainable degree of epidemic control.

Our study demonstrates that knowledge of all three key disease characteristics together, rather than any individual or pair of characteristics, is generally necessary for understanding disease controllability. Together, the effects of a large *R*_0_, short *τ*_*L*_, and large NSTI can compound with one another to diminish controllability to a greater degree than what would be expected from their individual effects. In this sense, the Omicron COVID-19 variant is a “perfect storm” disease that uniquely combines extreme values of all three factors. Consequently, this disease is far less controllable than any other in our study, including other COVID-19 variants. Even diseases like measles that combine only two factors from the set of large *R*_0_, short *τ*_*L*_, or large NSTI can prove extremely difficult if not impossible to control at practicably attainable resource levels. Conversely, the only disease in our study that lacks all three of these properties, SARS, is by far the most controllable and the only disease for which invasions into naive populations can be altogether prevented at low resource levels. It is perhaps no coincidence then that the 2002-2003 SARS outbreak was contained to 8098 known cases worldwide primarily with identification-isolation measures^29^ while in comparison, COVID-19 has spread to over 700 million total cases^17^. Our results suggest that this extreme difference in outcomes is not merely because COVID has much greater non-symptomatic transmission, but rather, is due to high levels of non-symptomatic transmission in conjunction with a moderately short latent period. Were COVID to have a longer latent period like hepatitis A, non-symptomatic transmission would be far less consequential. Together, these findings illustrate that due to interactions among disease parameters, controllability differences between two diseases may be much greater than one might intuitively expect based on differences in their individual characteristics. This result makes clear how modest changes the dynamic features of a novel disease may drive major changes in its epidemic trajectory, limiting opportunities to predict disease dynamics and shape public health responses based on prior experiences.

Interactions act as a double-edged sword for disease control. Just as the presence of one problematic disease characteristic can exacerbate the negative effects of the other two,its absence can also have a mitigating influence. Notably, reducing *R*_0_ has the added benefit of reducing the compounding impact of short latent periods and high levels of non-symptomatic transmission. A testing-isolation program may therefore see unexpectedly large gains in controllability when implemented in conjunction with an *R*_0_ reduction strategy, thereby allowing outbreaks to be curtailed at lower testing capacities than otherwise possible. This result underscores the critical importance of individual behavior in preventing wide-scale invasions of novel diseases. Although little can be done about a disease’s latent period and its degree of non-symptomatic transmission, its *R*_0_ can be effectively lowered through transmission-reduction measures, such as mask wearing^30^, social distancing^31^, and hand washing^32^. If widely adopted, these individual behaviors can have a disproportionately positive impact on controllability, in effect partially compensating for both supply shortfalls and problematic disease characteristics otherwise inaccessible to direct intervention. For example, even modest reductions in *R*_0_ could allow the original COVID-19 strain and hepatitis A to be contained with fewer than 10 test per thousand per day, while smallpox could be contained with fewer than 5 test per thousand per day (Extended Data Fig. 2a). These thresholds were obtained by a small number of countries within the first year of the COVID-19 pandemic^17^, so such rates may be achievable with increased effort to raise testing capacities early in the course of future epidemics. Wide-scale compliance with transmission-reducing behavior thus forms an essential component of successful out-break suppression under real-world resource constraints. Our results emphasize that this feature is not unique to COVID-19: across a broad spectrum of disease traits, epidemic containment typically requires adoption of testing-isolation strategies in conjunction with substantive changes in individual behavior.

Our study provides qualitative and definitive guidance for managing future novel disease outbreaks at the low levels of testing capacity expected during the initial phases of an epidemic. Testing-isolation strategies may protect the public from wide-scale disease spread without changes in individual behavior only for diseases like SARS, which lack large basic reproduction numbers, short latent periods, and moderate to substantial degrees of non-symptomatic transmission. For a disease that carries any one of these three properties, like smallpox or hepatitis A, outbreak suppression without wide-scale behavior change is possible but dubious. Epidemic containment will require rapid increases of testing capacity to a bare minimum of 10 tests per thousand individuals per day, although the actual number required may be much larger, especially in the face of real-world limitations like imperfect test accuracy and imperfect isolation. For diseases that exhibit two or more of these three properties, outbreak suppression without public compliance in transmission-reducing behavior is most likely impossible. Diseases with such properties are the most dangerous in terms of pandemic potential, and public health agencies should be on exceptionally high alert upon their emergence. Curtailing the invasion of such a disease in the absence of vaccinations is a feasible objective only if identification-isolation strategies are implemented in conjunction with broad public awareness of and adherence to precautionary measures like social distancing, masking, and personal hygiene measures.

## Methods

### Controlled reproduction number

The controlled disease reproduction number *R*_*c*_ is defined as the average number of secondary cases generated by an initial infection in an otherwise susceptible population partially protected by disease intervention measures^21^ (testing and isolation measures specifically for our study). This setting describes a scenario where government or health agencies attempt to mitigate the spread of an invading novel disease into a vulnerable population with no previously developed treatments or immunities. The value *R*_*c*_ = 1 represents a threshold for disease invasion into a naive population; when *R*_*c*_ > 1, an initial case introduced to an otherwise susceptible disease-free system will typically multiply and develop into an exponentially growing epidemic, while when *R*_*c*_ < 1, disease levels tend to rapidly die out. Reducing *R*_*c*_ to or below 1 using intervention measures thus provides a quantifiable control goal for health agencies seeking to suppress the spread of an invading disease.

We derive a mathematical expression for *R*_*c*_ using the next-generation operator method^33,34^ applied to a partial integro-differential equation compartmental model capable of representing a wide array of disease systems subject to resource-constrained testing and isolation controls^20^ (Supplementary Information). The expression is:

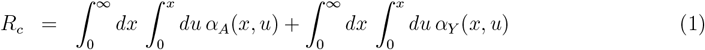

where we define the following notation:

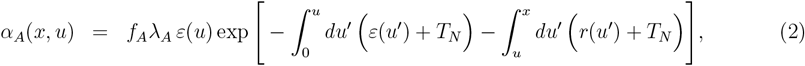

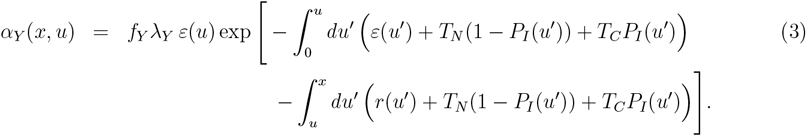

The first double integral in *R*_*c*_ represents the contribution from individuals who either remain asymptomatic or show only very mild symptoms throughout the course of an infection (denoted by the subscript ‘A’), while the second double integral represents contributions from individuals who eventually become moderately to severely symptomatic before recovering (denoted by the subscript ‘Y’). The parameter *f*_*A*_ denotes the fraction of all cases which remain mild or totally asymptomatic with *λ*_*A*_ representing the corresponding transmission of infected individuals (for some diseases, *f*_*A*_ = 0 for). The parameters *f*_*Y*_ and *λ*_*Y*_ are defined analogously for cases that eventually show moderate to severe symptoms. The functions *ε*(*x*) and *r*(*x*) denote the rates at which individuals transition from the latent state (meaning infected but not yet infectious) to the infectious state, and from the infectious state to the recovered state, respectively, which are dependent on the number of days *x* that an individual has been infected. Both rates are defined as hazard rate functions for the latent period distribution *f*_*ε*_(*x*) and recovery day distribution *f*_*r*_(*x*) as follows:

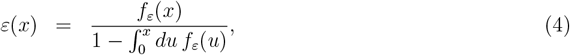

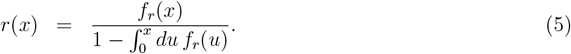

Symptom onset is encoded into the model by the function *P*_*I*_(*x*) denoting the probability that an infected individual in the ‘Y’ class will have begun to show symptoms by at least infection day *x*. Mathematically, this quantity is defined the cumulative distribution function of the incubation period distribution *f*_*I*_(*x*):

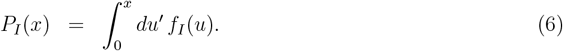

To clarify, *f*_*ε*_(*x*)*dx* is the probability for an infected individual to initially become contagious between infection days *x* and *x* + *dx* for some infinitesimal duration *dx, f*_*I*_(*x*)*dx* is the probability for an infected individual in the ‘Y’ class to begin showing symptoms between infection days *x* and *x* + *dx*, and *f*_*r*_(*x*)*dx* is the probability for an infected individual to recover between infection days *x* and *x* + *dx*.

Identification-isolation control strategies enter the model through the non-clinical *T*_*N*_ and clinical *T*_*C*_ testing expressions that target non-symptomatic and symptomatic individuals, respectively. These terms represent the rates at which individuals are tested, identified if infected, and subsequently transferred to an isolated state where they can no longer generate secondary infections. The mathematical form of these expressions is adopted from earlier works of ours^20,35^ (Supplementary Information). Resource constraints enter the model through a testing capacity *C* representing the maximum number of tests per-capita that can be administered and processed per day. Resource management decisions are modeled by allowing a fraction *ρ* of the testing capacity to be allocated to non-clinical testing and the reaming 1 − *ρ* allocated to clinical testing. In particular, optimal *ρ* values for reducing *R*_*c*_ can be chosen if known. The testing expressions *T*_*N*_ and *T*_*C*_ are increasing functions of their allocated testing capacities, decreasing to zero as capacity goes to zero, growing linearly with *C* at small but non-zero capacities, and saturating to a fixed finite constant as capacity approaches infinity. This limiting constant represents the average amount of time needed for a individual to acquire a test and receive results without delays or backlogs due to other patients, multiplied by the fraction of the population open to and accessible by testing efforts. Testing expressions also contain a number of other parameters that allow the model to tune between a low-efficacy random testing program or high-efficacy testing initiatives incorporating factors like contact tracing. The full mathematical details of these expressions as well as the underlying partial integro-differential equation disease model appear in the Supplementary Information.

### Key disease characteristics

We describe the natural (i.e., not subject to control interventions) properties of disease in terms of three key characteristics: the uncontrolled basic reproduction number *R*_0_, the mean latent period *τ*_*L*_, and the non-symptomatic transmission index NSTI. These quantities are defined mathematically as follows:

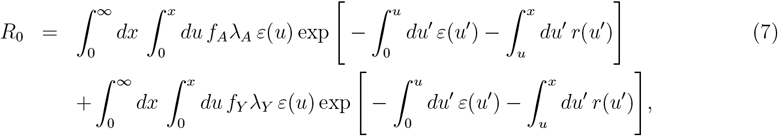

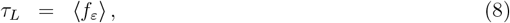

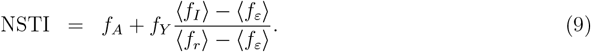

The expression for *R*_0_ is equivalent to *R*_*c*_ in Eq. (1) when the testing expressions *T*_*N*_ and *T*_*C*_ are set to zero. This quantity represents the expected number of new cases that will be generated by a single infected individual if they fail to be identified and isolated by control measures, or equivalently, if control measures are not active. The mean latent period *τ*_*L*_ is simply the expectation value of the latent period distribution *f*_*ε*_. NSTI is the sum of the fraction of cases that will never develop moderate to severe symptoms plus the fraction of cases that will develop such symptoms modulated by the incubation-latent offset factor ⟨*f*_*I*_⟩ − ⟨*f*_*ε*_⟩ */* ⟨*f*_*r*_⟩ − ⟨*f*_*ε*_⟩. The case ⟨*f*_*I*_⟩ > ⟨*f*_*ε*_⟩ (mean incubation period greater than mean latent period) represents a disease for which pre-symptomatic transmission occurs, where the incubation-latent offset factor is equivalent to the fraction of the infectious period for which symptoms are not present. Here, NSTI combines pre- and asymptomatic transmission together to give an overall measure of the level of non-symptomatic transmission in a disease. Conversely, the case ⟨*f*_*I*_⟩ < ⟨*f*_*ε*_⟩ represents a disease for which symptom onset typically occurs before substantial levels of infectiousness develop in an individual, and the magnitude of the incubation-latent offset factor gives the mean duration of this pre-contagious window of symptom presentation relative to the mean infectious period. The negative sign of the incubation-latent offset factor reduces NSTI and counteracts any asymptomatic transmission contribution from *f*_*A*_. Very small or negative values of NSTI thus indicate a disease for which non-symptomatic transmission is insignificant, and negative values in particular indicate a disease where symptom presentation aids in identifying infected individuals before they begin transmitting. We note that NSTI functions similarly to the *θ* parameter of Fraser et al.^18^ but contains additional information indicating the degree to which pre-infectiousness symptom onset aids the early identification of infected cases.

### Model parameterization

The parameters *f*_*A*_, *f*_*Y*_, *λ*_*A*_, and *λ*_*Y*_ in Eq. (1) and the distributions *f*_*ε*_, *f*_*r*_ and *f*_*I*_ in Eqs. (2), (3), and (4), respectively, are selected based on the results of literature searches for the original COVID-19 strain^24,28,36–43^, Delta COVID-19 variant^41–50^, Omicron COVID-19 variant^39,41–43,45,51–56^, SARS^18,19,39,57–62^, hepatitis A^19,63–66^, measles^58,67–71^, and smallpox^19,72–76^. Numerical estimates for these quantities, where available, are typically subject to large uncertainties that can vary widely across the epidemiological and mathematical modeling literature. We therefore consider entire ranges of values for the parameters found to have multiple reliable estimates, accounting for both reported values as well as confidence intervals and interquartile ranges. Parameters for which we found scant or no reliable estimates are assigned single values based on the limited data available, or are given assumed values if necessary. Ranges, values, and corresponding references used in our study are given in Extended Data Table 1.

Latent period *f*_*ε*_, incubation period *f*_*I*_, and recovery day *f*_*r*_ distributions are assumed to take the general form of gamma distributions with means and standard deviation taken from the literature when possible. For most diseases, we find ample estimates for mean latent and incubation period durations but few if any estimates for the standard deviations. Therefore, to achieve uniformity in the parameter sampling procedure among diseases, we consider ranges for the mean latent and incubation periods and single values for their standard deviations. Mean incubation periods are drawn relative to the mean latent period so that the mathematical model may directly incorporate differences between symptom and infectiousness onset reported in the literature. In both the smallpox and measles literature, it is often unclear whether the onset of observable symptoms allowing outward recognition of the disease, along with the corresponding reported incubation times, refers to the onset of the characteristic rash or to the onset of the prodromal flu-like phase. The ranges selected for ⟨*f*_*ε*_⟩ and ⟨*f*_*I*_⟩ account for both possibilities. For smallpox, we also find a lack of consensus in the literature regarding the degree of infectiousness, if any, during the prodromal phase; our selected range for ⟨*f*_*ε*_⟩ along with allowing ⟨*f*_*I*_⟩ to be both larger and smaller than ⟨*f*_*ε*_⟩ accounts for both viewpoints. For all diseases, we are unaware of any direct estimates for the recovery day distribution *f*_*r*_(*x*) or the distribution of infectious period lengths. In the literature, infectiousness is often estimated indirectly from the CT values of PCR tests or the results of viral isolation studies. Where possible, we select values for ⟨*f*_*r*_⟩ using reported time windows of live viral isolation as a proxy for the mean contagious period, considering also the results of mathematical modeling studies and more qualitative or general statements regarding the timing and duration of infectiousness. Depending on the context and phrasing of the literature, the total duration of infection ⟨*f*_*r*_⟩ is written using the mean contagious period duration relative to either symptom onset or infectiousness onset.

For diseases having mild or totally asymptomatic cases in addition to symptomatic cases, the fraction *f*_*A*_ is subject to much uncertainty, potentially varying based on demographics, under reporting, and the precise meaning of a ‘mild’ case. We thus consider large *f*_*A*_ ranges around reported values and set *f*_*Y*_ = 1 − *f*_*A*_. Likewise, for diseases with reported differences in transmissibility between severe and mild or asymptomatic cases, the relative degree of transmissibility is highly uncertain, so we set *λ*_*Y*_ = *λλ*_*A*_ and assume large ranges for the relative transmissibility *λ* based on limited data available. The final parameter *λ*_*A*_ is determined after all other parameters have been selected by first drawing a value for the uncontrolled basic reproduction number *R*_0_ from a range determined from the literature, and then scaling *λ*_*A*_ such that Eq. (7) equals the selected *R*_0_ value.

### Numerical method for controllability threshold testing capacity

From the parameter ranges in Extended Data Table 1, we select 6000 parameter sets for each disease using Latin hypercube sampling, assuming uniform distributions over each parameter range. Each of these sets are then input into the expression for *R*_*c*_ in Eq. (1), and the minimum testing capacity required to reduce *R*_*c*_ to 1 is subsequently found using a two step optimization process. First, we utilize the *fmincon* function in *MatlabR2022b* to find the optimal allocation *ρ* of a given testing capacity *C* between non-clinical *T*_*N*_ and clinical *T*_*C*_ testing rates that minimizes *R*_*c*_. Next, we utilize *Matlab’s fzero* function to solve *R*_*c*_ −1 = 0 for the testing capacity *C* assuming the optimal allocation strategy *ρ* calculated in the first step. The result yields the minimum testing capacity required to reduce *R*_*c*_ to 1, i.e., the controllability threshold testing capacity. Data sets for the drawn parameter values and corresponding threshold testing capacities are provided for each disease as Supplementary Information.

### Multiple linear regression and marginal controllability costs

The full interaction multiple linear regression model for controllability threshold testing capacity *C*^*th*^ with independent variables *R*_0_, *τ*_*L*_, and NSTI takes the following form:

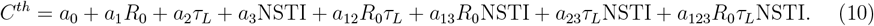

The constants *a*_12_, *a*_13_, and *a*_23_ represent the *R*_0_ : *τ*_*L*_, *R*_0_ : NSTI, and *τ*_*L*_ : NSTI two-way interactions, respectively, while *a*_123_ represents the three-way interaction. The two-way interaction model is obtained by setting *a*_123_ = 0 and the non-interacting model is obtained by further setting *a*_12_ = *a*_12_ = *a*_23_ = 0. Values for the regression coefficients are obtained using linear least squares fitting (Extended Data Table 2).

Marginal controllability costs of *R*_0_ increase, *τ*_*L*_ decrease, and NSTI increase are denoted 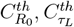, and 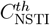, respectively. Mathematical expressions for these quantities are obtained from partial derivatives of *C*^*th*^ as follows:

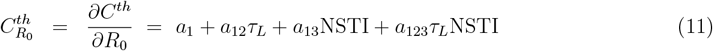

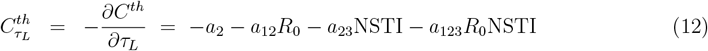

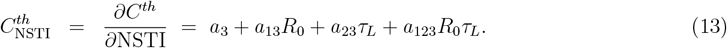

The negative sign in 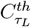 appears because we are considering the marginal cost of a latent period decrease rather than an increase. The above expression shows that marginal controllability costs only vary in response to the interaction terms. Thus, the functional behavior of marginal costs provide direct insight into the behavior of interactions and their influences on disease controllability.

## Data Availability

Parameter data sets drawn for each disease along with the corresponding calculated threshold testing capacities are provided as Supplementary Information.

## Acknowledgments

This work was funded by the Center of Advanced Systems Understanding (CASUS) which is financed by Germany’s Federal Ministry of Education and Research (BMBF) and by the Saxon Ministry for Science, Culture and Tourism (SMWK) with tax funds on the basis of the budget approved by the Saxon State Parliament. The University of Maryland supported the efforts of SP and WFF.

## Author Contributions

J.D. conceived the study, built the model, wrote code, collected data, interpreted results, and wrote the manuscript. S.P. wrote code, collected data, interpreted results, and revised the manuscript. J.M.C. and W.F.F. interpreted results, revised the manuscript, and supervised the project.

## Extended data

**Extended Data Fig. 1:**
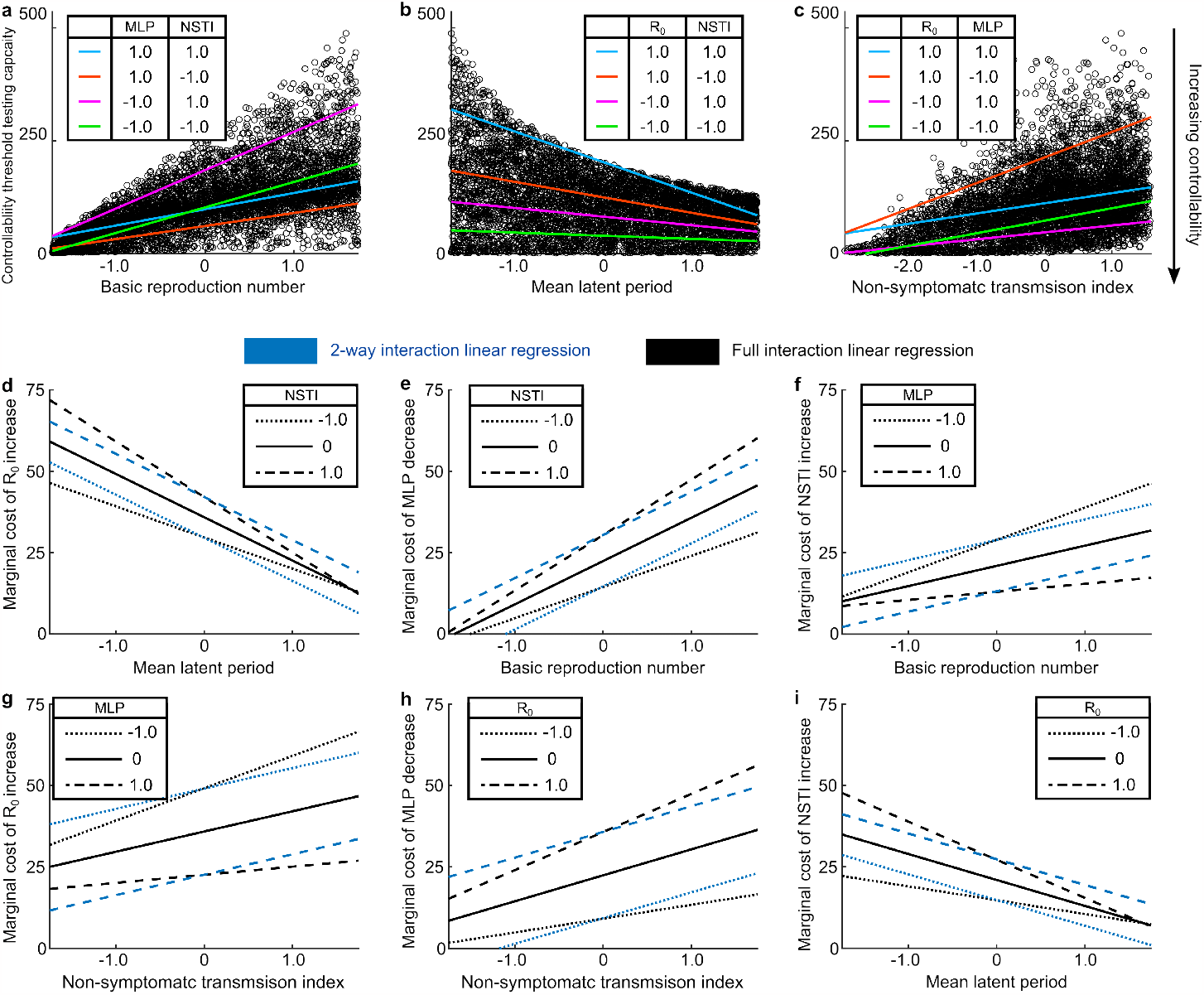
Controllability threshold testing capacities, linear regression fits, and marginal costs for a generically parameterized disease. Standardized units adopted for the basic reproduction number *R*_0_ (mean 8.25, SD 3.90), mean latent period MLP (mean 6.00 days, SD 1.73 days), and NSTI (mean 0.51, SD 0.23). Testing capacities and marginal costs given in tests per thousand per day. **a-c**, Threshold testing capacity scatter plot results as a function of key disease characteristics overlaid by fits from the full interaction liner regression model. **d-e**, Marginal controllability cost of key disease characteristics. Blue lines indicate marginal controllability cost calculated from the two-way interaction regression model, while black lines indicate the full two and three-way interaction model. Differences between corresponding blue and black lines indicate influences of the three-way interaction term.

**Extended Data Fig. 2:**
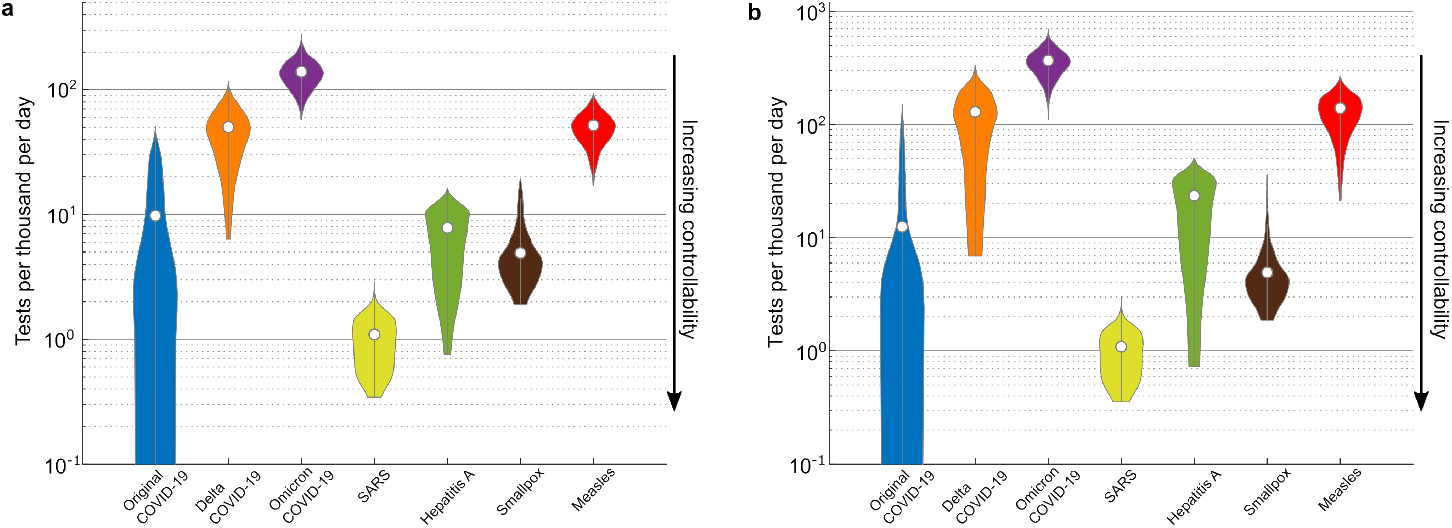
Controllability threshold testing capacities for testing-isolation interventions combined with transmission reduction measures (i.e. social distancing). Violin plots of controllability thresholds for 6000 points sampled from the parameter spaces of each disease, where white dots indicate median controllability levels. Basic reproduction number values are sampled from the ranges in Extended Data Table 1 and then reduced by a factor of 1/3 to simulate adoption of contact rate or transmissibility reducing measures like social distancing and masking, respectively. Log-scaled y-axes in all plots indicate threshold testing capacities for suppressing or preventing disease outbreaks in units of tests administered and processed per thousand individuals per day. Smaller thresholds indicate greater degrees of controllability. **a**, Optimistic high-efficacy contact tracing testing scenario considered in the main text. **b**, Pessimistic low-efficacy random sampling testing scenario.

**Extended Data Fig. 3:**
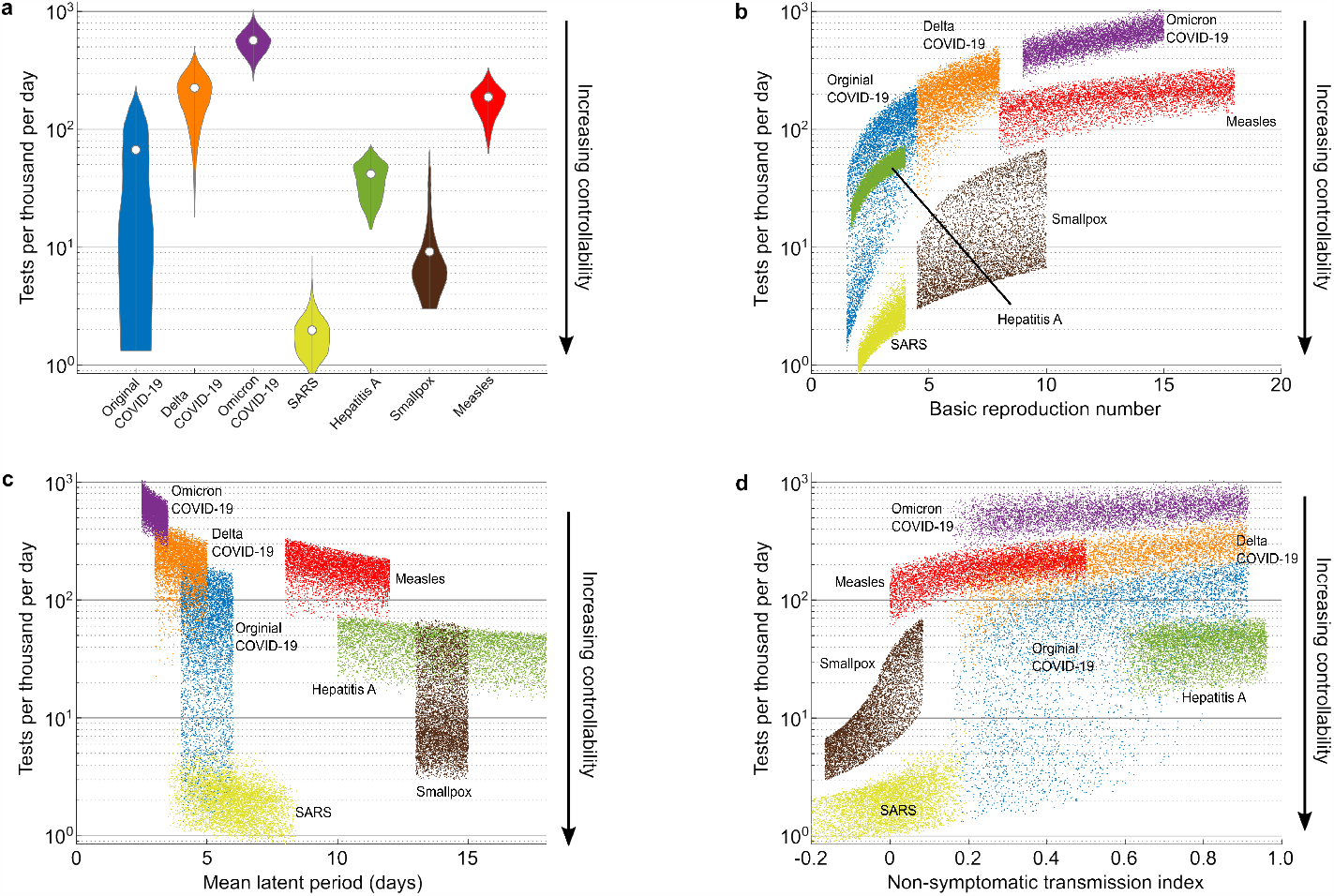
Controllability of seven past and current diseases under a pessimistic low-efficacy random sampling testing scenario. Log-scaled y-axes in all plots indicate threshold testing capacities for suppressing or preventing disease outbreaks in units of tests administered and processed per thousand individuals per day. Smaller thresholds indicate greater degrees of controllability in the sense that fewer testing resources are required to prevent large scale disease invasions. Thresholds are larger than those of the optimistic scenario given in Fig. 1 of the main text but otherwise follow similar trends. **a**, Violin plots of controllability thresholds for 6000 points sampled from the parameter spaces of each disease, where white dots indicate median controllability levels. **b – d**, Controllability thresholds plotted as a function of key disease characteristics for each disease, where each point represents a particular combination of parameters sampled from a disease’s parameter space.

**Extended Data Fig. 4:**
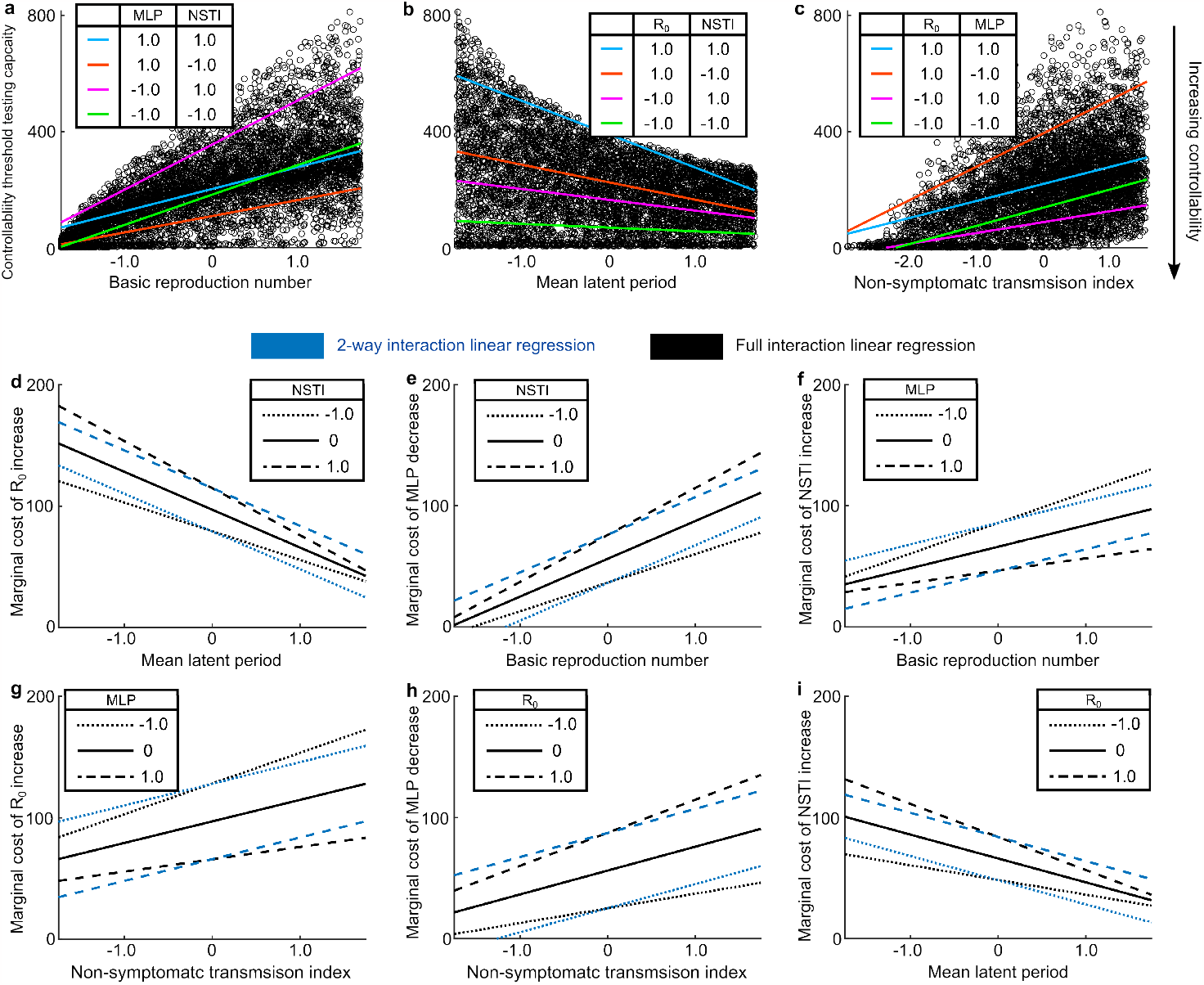
Controllability threshold testing capacities, linear regression fits, and marginal costs for a generically parameterized disease under a pessimistic low-efficacy random sampling testing scenario. Standardized units adopted for the basic reproduction number *R*_0_ (mean 8.25, SD 3.90), mean latent period MLP (mean 6.00 days, SD 1.73 days), and NSTI (mean 0.51, SD 0.23). Testing capacities and marginal costs given in tests per thousand per day. Thresholds and marginal costs are generally larger than those of the optimistic scenario given in Extended Data Fig. 1 but otherwise behave analogously. **a-c**, Threshold testing capacity scatter plot results as a function of key disease characteristics overlaid by fits from the full interaction liner regression model. **d-e**, Marginal controllability cost of key disease characteristics. Blue lines indicate marginal controllability cost calculated from the two-way interaction regression model, while black lines indicate the full two and three-way interaction model. Differences between corresponding blue and black lines indicate influences of the three-way interaction term.

**Extended Data Table 1:** Parameter ranges and values for modeled diseases

| Disease | Basic reproduction number $R_0$ | Latent period distribution $f_\varepsilon$ (days)* | Incubation period distribution $f_I$ (days)* | Recovery day distribution $f_r$ (days)* | Fraction asymptomatic $f_A$ | Relative symptomatic transmissibility $\lambda$ |
| --- | --- | --- | --- | --- | --- | --- |
| COVID-19 Original variant | [1.5, 4.5] | $\langle f_\varepsilon \rangle \in [4.0, 6.0]$<br>$\sigma = 2.72$ | $\langle f_I \rangle \in \langle f_\varepsilon \rangle + [0.5, 2.0]$<br>$\sigma = 2.22$ | $\langle f_r \rangle = \langle f_I \rangle + 9.0$<br>$\sigma = 1.00$ | [0.10, 0.90] | [1.0, 3.0] |
| COVID-19 Delta variant | [4.5, 8.0] | $\langle f_\varepsilon \rangle \in [3.0, 5.0]$<br>$\sigma = 2.40$ | $\langle f_I \rangle \in \langle f_\varepsilon \rangle + [0.5, 2.0]$<br>$\sigma = 2.90$ | $\langle f_r \rangle = \langle f_I \rangle + 9.0$<br>$\sigma = 1.00$ | [0.10, 0.90] | [1.0, 3.0] |
| COVID-19 Omicron variant | [9.0, 15.0] | $\langle f_\varepsilon \rangle \in [2.5, 3.5]$<br>$\sigma = 1.49$ | $\langle f_I \rangle \in \langle f_\varepsilon \rangle + [0.5, 2.0]$<br>$\sigma = 1.32$ | $\langle f_r \rangle = \langle f_I \rangle + 9.0$<br>$\sigma = 1.00$ | [0.10, 0.90] | [1.0, 3.0] |
| SARS | [2.0, 4.0] | $\langle f_\varepsilon \rangle \in [5.5, 7.5]$<br>$\sigma = 1.00$ | $\langle f_I \rangle \in \langle f_\varepsilon \rangle + [-2.0, 0]$<br>$\sigma = 2.62$ | $\langle f_r \rangle = \langle f_I \rangle + 16.0$<br>$\sigma = 1.00$ | [0, 0.20] | [1.0, 3.0] |
| Hepatitis A | [1.70, 4.0] | $\langle f_\varepsilon \rangle \in [10.0, 18.0]$<br>$\sigma = 1.00$ | $\langle f_I \rangle \in \langle f_\varepsilon \rangle + [11.0, 18.0]$<br>$\sigma = 1.00$ | $\langle f_r \rangle = \langle f_I \rangle + 10.0$<br>$\sigma = 1.00$ | [0.10, 0.90] | 1.0 |
| Measles | [8.0, 18.0] | $\langle f_\varepsilon \rangle \in [8.0, 12.0]$<br>$\sigma = 1.00$ | $\langle f_I \rangle \in \langle f_\varepsilon \rangle + [0, 4.0]$<br>$\sigma = 1.00$ | $\langle f_r \rangle = \langle f_\varepsilon \rangle + 8.0$<br>$\sigma = 1.00$ | 0 | 1.0 |
| Smallpox | [4.5, 10.0] | $\langle f_\varepsilon \rangle \in [13.0, 15.0]$<br>$\sigma = 1.00$ | $\langle f_I \rangle \in \langle f_\varepsilon \rangle + [-2.0, 1.0]$<br>$\sigma = 1.00$ | $\langle f_r \rangle = \langle f_\varepsilon \rangle + 12.0$<br>$\sigma = 1.00$ | 0 | 1.0 |
| Generically parameterized disease | [1.5, 15.0] | $\langle f_\varepsilon \rangle \in [3.0, 9.0]$<br>$\sigma = 1.00$ | $\langle f_I \rangle \in \langle f_\varepsilon \rangle + [-3.0, 5.0]$<br>$\sigma = 1.00$ | $\langle f_r \rangle = \langle f_\varepsilon \rangle + 12.0$<br>$\sigma = 1.00$ | [0, 0.90] | [1.0, 3.0] |

\* Assumed gamma distributions with means $\langle \dots \rangle$ and standard deviations $\sigma$ in units of days

**Extended Data Table 2:** References for parameter ranges and values

| Disease | Basic reproduction number $R_0$ | Latent period distribution $f_\varepsilon^*$ | Incubation period distribution $f_I^*$ | Recovery day distribution $f_r^*$ | Fraction asymptomatic $f_A$ | Relative symptomatic transmissibility $\lambda$ |
| --- | --- | --- | --- | --- | --- | --- |
| COVID-19 Original strain | 24,36 | $\langle f_\varepsilon \rangle^{28,37}, \sigma^{28}$ | $\langle f_I \rangle^{28,38}, \sigma^{38}$ | $\langle f_r \rangle^{39,40}, \sigma^\dagger$ | 41,42 | 42,43 |
| COVID-19 Delta variant | 44,45 | $\langle f_\varepsilon \rangle^{46,47}, \sigma^{46}$ | $\langle f_I \rangle^{46,48,49}, \sigma^{46}$ | $\langle f_r \rangle^{46,50}, \sigma^\dagger$ | 41,42 | 42,43 |
| COVID-19 Omicron variant | 45,51 | $\langle f_\varepsilon \rangle^{52,53}, \sigma^{53}$ | $\langle f_I \rangle^{53-55}, \sigma^{53}$ | $\langle f_r \rangle^{39,56}, \sigma^\dagger$ | 41,42 | 42,43 |
| SARS | 18,57 | $\langle f_\varepsilon \rangle^{19,39}, \sigma^\dagger$ | $\langle f_I \rangle^{58-61}, \sigma^{60}$ | $\langle f_r \rangle^{19,39,60}, \sigma^\dagger$ | 39,62 | † |
| Hepatitis A | 63,64 | $\langle f_\varepsilon \rangle^{63}, \sigma^\dagger$ | $\langle f_I \rangle^{19,63,65}, \sigma^\dagger$ | $\langle f_r \rangle^{65,66}, \sigma^\dagger$ | 63,66 | † |
| Measles | 67,68 | $\langle f_\varepsilon \rangle^{58,69-71}, \sigma^\dagger$ | $\langle f_I \rangle^{58,70}, \sigma^\dagger$ | $\langle f_r \rangle^{58,71}, \sigma^\dagger$ | † | † |
| Smallpox | 19,72 | $\langle f_\varepsilon \rangle^{73-75}, \sigma^\dagger$ | $\langle f_I \rangle^{19,73}, \sigma^\dagger$ | $\langle f_r \rangle^{73,75,76}, \sigma^\dagger$ | † | † |
| Generically parameterized disease | † | $\langle f_\varepsilon \rangle^\dagger, \sigma^\dagger$ | $\langle f_I \rangle^\dagger, \sigma^\dagger$ | $\langle f_r \rangle^\dagger, \sigma^\dagger$ | † | † |

\* References given for means $\langle \dots \rangle$ and standard deviations $\sigma$ of distributions
† Assumed value

**Extended Data Table 3:**
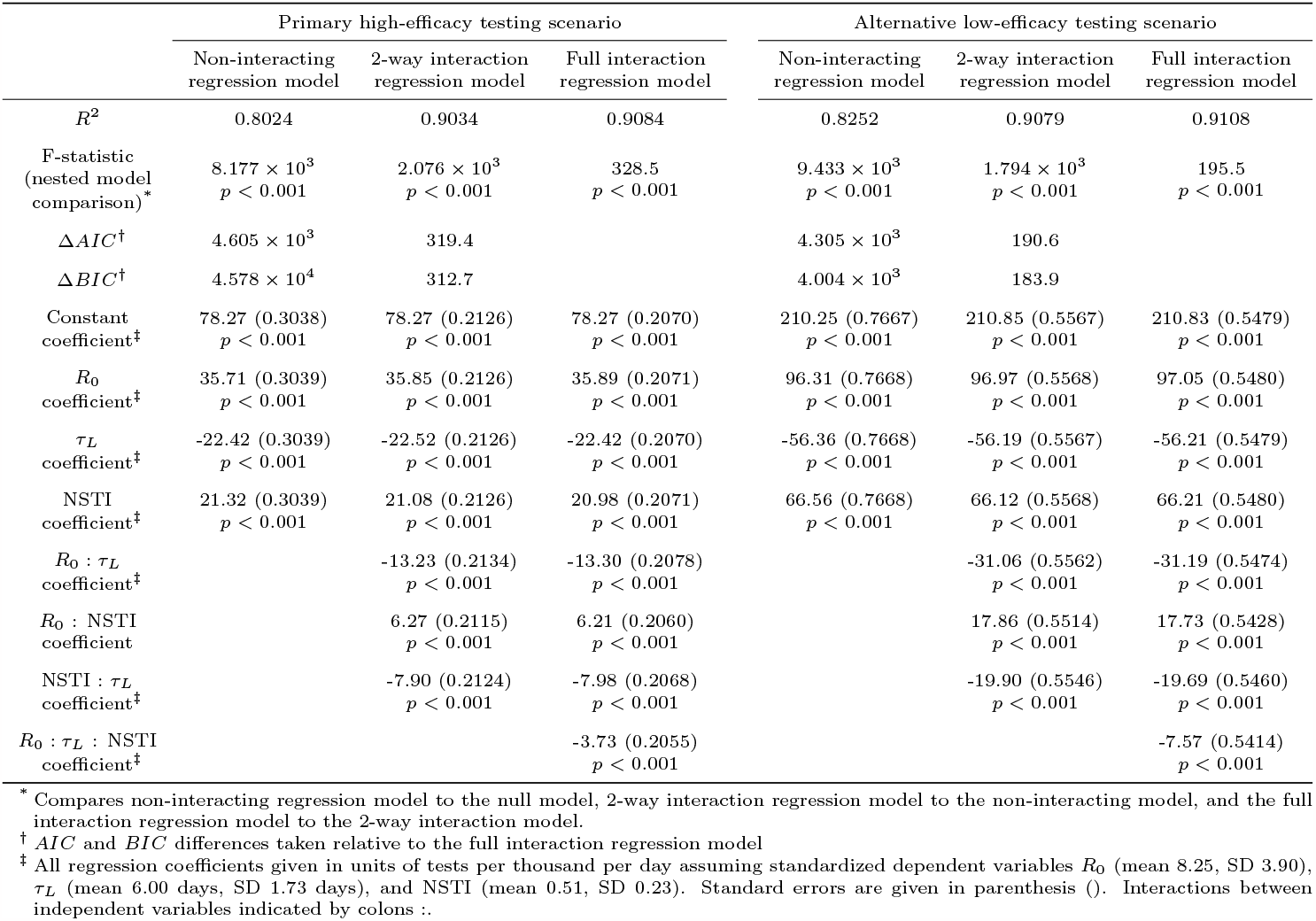
Multi-linear regression coefficients for generically parameterized disease

