## Supplementary Information for "Testing-isolation interventions will likely be insufficient to contain future novel disease outbreaks"

### Supplementary Equations

#### Non-linear disease model description

The non-linear partial integro-differential equation system underlying our study follows a SEIR compartmental structure that utilizes the age of infection (i.e., the number of days an individual has been infected) to decouple symptom status from any one model compartment. This unique feature allows our model to represent a wide variety of diseases with differing relationships between symptom and infectiousness onset all under a single analytical framework. This system given here has been adapted from previous iterations of this model structure; see Calabrese and Demers (2022)[35] and Demers et al. (2023)[20] for previous model analyses and further in-depth reasoning behind the mathematical formulation given here.

##### Uncontrolled disease dynamics

Our underlying disease model describes a homogeneously mixed population of $N$ individuals partitioned into susceptible, exposed, infectious, and recovered classes based on their state of disease.

Exposed and infectious classes are further partitioned into those who will eventually show moderate to severe symptoms over the course of the disease and those who will remain permanently asymptomatic or only develop mild symptoms. These symptom-based partitions are referred to as eventually symptomatic (ES) and permanently asymptomatic (PA), respectively. Model equations describe how the total number of individuals in each class change over time $t$.

Individuals begin in the susceptible class $S(t)$ and may become infected through contact with the infectious classes. Upon infection, individuals enter either the PA exposed class $E_A(t)$ or the ES exposed class $E_Y(t)$ with probabilities $f_A$ and $f_Y$, respectively. While in an exposed class, infected individuals are not contagious and cannot transmit the disease to others. Exposed individuals next transition to their respective ES or PA infectious classes, $Y(t)$ and $A(t)$, with transition rates $\varepsilon(x)$ dependent on how long $x$ one has been infected, i.e., the age of infection (those who have been infected for longer are more likely to be the next ones to transition). We therefore continuously index the ES exposed class by the age of infection $x$ and let $e_Y(t,x)$ be the distribution of $E_Y(t)$ over all infection ages, meaning that $e_A(t,x)dx$ give the number of individuals in $E_Y(t)$ who have been infected for between $x$ and $x+dx$ infection days that $E_Y(t) = \int_0^\infty dx\, e_Y(t,x)$. The exposed PA class $E_A(t,x)$ as well as the infectious ES $Y(t)$ and PA $A(t)$ are likewise indexed as $e_A(x,t), y(x,t)$, and $a(x,t)$, respectively. Infectious individuals subsequently transition to the recovered class $R(t)$ at age of infection dependent recovery rate $r(x)$. Recovered individuals are no longer infectious and are assumed to have attained permanent immunity (relaxing the permanent immunity assumption does not alter the expressions for $R_0$ or $R_c$ given in the Methods of the main text). The model equations governing this disease system are as follows:

$$\dot{S}(t) = -\lambda_A \frac{A(t)}{N} S(t) - \lambda_Y \frac{Y(t)}{N} S(t) \tag{S1a}$$

$$\partial_t e_A(t,x) + \partial_x e_A(t,x) = -\varepsilon(x)\, e_A(t,x) \tag{S1b}$$

$$e_A(t,0) = f_A \left( \lambda_A \frac{A(t)}{N} S(t) + \lambda_Y \frac{Y(t)}{N} S(t) \right) \tag{S1c}$$

$$\partial_t e_Y(t,x) + \partial_x e_Y(t,x) = -\varepsilon(x)\, e_Y(t,x) \tag{S1d}$$

$$e_Y(t,0) = f_Y \left( \lambda_A \frac{A(t)}{N} S(t) + \lambda_Y \frac{Y(t)}{N} S(t) \right) \tag{S1e}$$

$$\partial_t a(t,x) + \partial_x a(t,x) = \varepsilon(x)\, e_A(t,x) - r(x) a(t,x) \tag{S1f}$$

$$a(t,0) = 0 \tag{S1g}$$

$$\partial_t y(t,x) + \partial_x y(t,x) = \varepsilon(x)\, e_Y(t,x) - r(x) y(t,x) \tag{S1h}$$

$$y(t,0) = 0 \tag{S1i}$$

$$\dot{R}(t) = \int_0^\infty dx\, r(x) \Big( a(t,x) + y(t,x) \Big) \tag{S1j}$$

$$E_A(t) = \textstyle\int_0^\infty dx e_A(t,x), \quad E_Y(t) = \int_0^\infty dx\, e_Y(t,x), \quad A(t) = \int_0^\infty dx\, a(t,x), \quad Y(t) = \int_0^\infty dx\, y(t,x) \tag{S1k}$$

Overdots in the above equations denote ordinary derivatives with respect to time $t$. The parameter $N$ denotes the total population size and $\lambda_A$ and $\lambda_Y$ denote the transmission rates for the PA and ES infectious classes, respectively. The transition rates $\varepsilon(x)$ and $r(x)$ are determined from latent and recovery day distributions, respectively, as indicated in Eqs. (4) and (5) of the Methods section in the main text. Rates of new infections (i.e., individuals with infection age $x = 0$) entering the PA and ES exposed classes are given by the boundary terms $e_A(t, 0)$ and $e_Y(t, 0)$, respectively. The boundary terms $a(t, 0) = y(t, 0) = 0$ indicate that no individuals can become instantaneously contagious upon infection (they must first pass through the non-infectious exposed class). The integrals in Eq. (S1k) relate the total sizes of the infected classes to their corresponding distributions.

In the above disease system, no one model compartment is deemed to represent a symptomatic state. Rather, our model assumes that a portion $P_I(x)$ of the $E_Y$ and $Y$ compartments will have begun showing symptoms by infection day $x$. The function $P_I(x)$ is equivalent to the probability that any one infected ES individual will develop symptoms on or before the $x^{th}$ infection day, calculated as the cumulative distribution function of disease incubation period distribution (Eq. (6) of the Methods section in the main text). The symptomatic $X_S(t)$ and non-symptomatic $X_N(t)$ infected population sizes at time $t$ are given as follows:

$$X_S(t) = \int_0^\infty dx\, P_I(x)\Big(e_Y(t,x) + y(t,x)\Big), \tag{S2}$$

$$X_N(t) = E_A(t) + A(t) + \int_0^\infty dx\, \Big(1 - P_I(x)\Big)\Big(e_Y(t,x) + y(t,x)\Big). \tag{S3}$$

##### Resource constrained testing formulation

Our testing model assumes that due to resource limitations, there exists a maximum rate at which tests can be administered to a population and subsequently processed to deliver results. The testing capacity $C$ denotes this maximum rate per capita. Testing resources are allocated to two types of testing, clinical and non-clinical, which act to identify symptomatic and mild to non-symptomatic infected individuals, respectively. Non-clinical resources are accessible to the general population through initiatives like public testing centers or workplace monitoring programs while clinical resources are reserved for individuals showing sufficiently strong symptoms at hospitals or health centers. Different choices of allocation strategy allow different fractions of the available resources to be reserved for the most severely ill cases. These types of resource management strategies were recommended, for example, by public health agencies faced with extreme testing supply shortfalls amid the initial COVID-19 pandemic. We define the allocation parameter $\rho$ such that $\rho C$ testing resources are allocated to non-clinical testing with the remainder $(1 - \rho)C$ are reserved for clinical testing.

The average time a test-seeking individual must wait to actually acquire a test and receive results depends on both the resource supply and the number of people actively seeking testing, i.e the testing demand. We assume the following forms of the non-clinical $\tau_N$ and clinical $\tau_C$ mean waiting times:

$$\tau_N = \tau + \frac{\kappa_N\Big(X_N(t) + \big(1-\eta_N\big)\big(S(t)+R(t)\big)\Big)}{\rho C N} \tag{S4}$$

$$\tau_C = \tau + \frac{\kappa_C\Big(X_S(t) + \big(1-\eta_C\big)\big(S(t)+R(t)\big)\Big)}{\big(1-\rho\big) C N}. \tag{S5}$$

In the above expressions, $\tau$ is the mean waiting time for acquiring testing and receiving results without any backlogs or delays due to other patients. We set this value equal to 1 day in our simulations. The non-clinical testing demand is given by the numerator of the fraction term in Eq. (S4), comprising a portion $\kappa_N$ of the non- or mildly symptomatic infected population and a portion $\kappa_N\big(1-\eta_N\big)$ of the uninfected population. Here, $\eta_N \in [0,1)$ is the non-clinical concentration parameter. This term accounts for biases that can make testing more likely to be applied to infected rather than uninfected individuals. The case $\eta_N = 0$ represents a purely random population monitoring program with no biases, where any one individual is equally likely as another to be tested . Biases that increase $\eta_N$ may exist, for example, by the fact that individuals with recent exposures, suspected recent exposures, or mild symptom presentation are more likely to be infected compared to a random member of the population, and these individuals are more likely to seek testing compared to those who have no explicit reason to do so. Contact tracing programs that actively seek out likely cases and encourage those individuals to seek testing are also a source of bias. Larger $\eta_N$ values correspond to greater shares of the limited available testing capacity being used on infected individuals (those who will test positive) over uninfected individuals (those who will test negative). Once testing results are received, test-positive individuals are identified as infected and are subsequently isolated to prevent further disease transmissions. Thus, larger $\eta_N$ values correspond to greater degrees of epidemic control efficacy achievable by non-clinical testing. The quantity $\kappa_N \in (0,1]$ is the non-clinical accessibility parameter denoting the fraction of the population open to and accessible by testing efforts. This factor may be less than 1 due to the distrust and non-compliance of some individuals in regards to public health initiatives, as well as the fact that some individuals may be geographically, socially, or communicatively isolated from the reach of testing supplies and contact tracers.

The mean clinical testing time is defined analogously to that of non-clinical testing, with clinical concentration parameter $\eta_C \in [0,1)$, clinical accessibility parameter $\kappa_C \in (0,1]$, and clinical testing demand comprising portions of the symptomatic infected population and uninfected population. Given that the presentation of moderate to severe visually identifiable symptoms is a pre-requisite to accessing clinical testing, the concentration $\eta_C$ is ideally equal to 1. More realistically, misdiagnoses, misuse of tests, and similar symptom presentation from other diseases may make $\eta_C$ slightly less than 1, although it should still be much greater than the $\eta_N$ of non-clinical testing. The clinical accessibility parameter likewise will be close if not equal to 1, assuming all individuals with moderate to severe symptoms will feel ill enough to seek medical attention. For our simulations, we set $\eta_C = 0.99$ and $\kappa_C = 1.00$. Non-clinical testing parameters are selected for low and high-efficacy cases based in part on our analysis in Calabrese and Demers (2022)[35]. For the low-efficacy case, non-clinical testing is a simple population-wide random monitoring program with $\eta_N = 0$ and $\kappa_N = 1.00$. For the high-efficacy case, we set $\eta_N = 0.70$ to represent a generous upper limit of what is practically achievable with a highly effective contact tracing program, with $\kappa_N = 0.75$ representing the logistical challenges of implementing such a program over a large population.

##### Testing and isolation disease dynamics

Testing and isolation strategies enter the disease model as transition rates from the infected classes to an isolated / quarantined class $Q(t)$ with infection age distribution $q(t,x)$. In the isolated class, individuals do not contact others, and are thus unable to spread disease, eventually recovering and returning to the general population at recovery rate $r(x)$. Testing and isolation strategies modify the system in Eq. (S1) as follows:

$$\begin{aligned}
\dot{S}(t) &= -\lambda_A \frac{A(t)}{N-Q(t)} S(t) - \lambda_Y \frac{Y(t)}{N-Q(t)} S(t) && \text{(S6a)}\\
\partial_t e_A(t,x) + \partial_x e_A(t,x) &= -\varepsilon(x)\, e_A(t,x) - \kappa_N \tau_N^{-1} e_A(t,x) && \text{(S6b)}\\
e_A(t,0) &= f_A \left( \lambda_A\, \beta \frac{A(t)}{N-Q(t)} S(t) + \lambda_Y\, \beta \frac{Y(t)}{N-Q(t)} S(t) \right) && \text{(S6c)}\\
\partial_t e_Y(t,x) + \partial_x e_Y(t,x) &= -\varepsilon(x)\, e_Y(t,x) - \kappa_C \tau_C^{-1} P_I(x) e_Y(t,x) && \text{(S6d)}\\
&\quad -\kappa_N \tau_N^{-1} \big(1 - P_I(x)\big) e_Y(t,x) &&\\
e_Y(t,0) &= f_Y \left( \lambda_A\, \beta \frac{A(t)}{N-Q(t)} S(t) + \lambda_Y\, \beta \frac{Y(t)}{N-Q(t)} S(t) \right) && \text{(S6e)}\\
\partial_t a(t,x) + \partial_x a(t,x) &= \varepsilon(x)\, e_A(t,x) - r(x) a(t,x) - \kappa_N \tau_N^{-1} a(t,x) && \text{(S6f)}\\
a(t,0) &= 0 && \text{(S6g)}\\
\partial_t y(t,x) + \partial_x y(t,x) &= \varepsilon(x)\, e_Y(t,x) - r(x) y(t,x) - \kappa_C \tau_C^{-1} P_I(x) y(t,x) && \text{(S6h)}\\
&\quad -\kappa_N \tau_N^{-1} \big(1 - P_I(x)\big) y(t,x) &&\\
y(t,0) &= 0 && \text{(S6i)}\\
\partial_t q(t,x) + \partial_x q(t,x) &= -r(x) q(t,x) + \kappa_C \tau_C^{-1} P_I(x) \Big[ e_Y(t,x) + y(t,x) \Big] && \text{(S6j)}\\
&\quad +\kappa_N \tau_N^{-1} \Big[ e_A(t,x) + a(t,x) + \big(1 - P_I(x)\big)\big(e_Y(t,x) + y(t,x)\big) \Big] &&\\
q(t,0) &= 0 && \text{(S6k)}\\
\dot{R}(t) &= \int_0^\infty dx\, r(x) \Big( a(t,x) + y(t,x) + q(t,x) \Big) && \text{(S6l)}
\end{aligned}$$

$$\begin{gathered}
E_A(t) = \textstyle\int_0^\infty dx e_A(t,x), \quad E_Y(t) = \int_0^\infty dx\, e_Y(t,x), \qquad \text{(S6m)}\\
A(t) = \textstyle\int_0^\infty dx\, a(t,x), \quad Y(t) = \int_0^\infty dx\, y(t,x), \quad Q(t) = \int_0^\infty dx\, q(t,x).
\end{gathered}$$

Note that in the above equations, we adopt the convention that $\tau_C^{-1}$ and $\tau_N^{-1}$ achieve their limiting value of 0 when $C = 0$, thus recovering the uncontrolled model (S1) when no testing resources are available. The terms $\kappa_N \tau_N^{-1}$ and $\kappa_C \tau_C^{-1}$ are the transition rates of non-symptomatic and symptomatic infected individuals, respectively, into the isolated class. When the clinical and non-clinical testing demands are far below the testing capacity, we have $\tau_N^{-1} \approx \tau_C^{-1} \approx \tau^{-1}$ with the total number of test administered and processed per day growing linearly with demand. As demands exceed the testing capacity and grow towards $\infty$, the rates $\tau_N^{-1}$ and $\tau_C^{-1}$ approach 0 such that the total number of tests administered processed per day (including those administered to non-infected individuals) approaches $CN$.

#### Controlled and basic reproduction numbers

The basic and controlled reproduction numbers for the systems (S1) and (S6), respectively, are calculated using the next-generation operator technique for compartmental models[17,18]. These quantities are calculated using the disease model dynamics linearized about the disease-free equilibrium, where the entire population is assumed to be susceptible. Reproduction numbers determine the local asymptotic stability of the disease-free state, being stable if reproduction numbers fall below 1 and becoming unstable as reproduction numbers grow beyond 1. Letting $T_N$ and $T_C$ denote the disease-free equilibrium evaluation of the transition rates $\kappa_N \tau_N^{-1}$ and $\kappa_C \tau_C^{-1}$, respectively, we find the following:

$$T_N = \begin{cases} 0, & C = 0 \text{ or } \rho = 0 \\ \kappa_N \left[ \tau + \kappa_N \frac{1-\eta_N}{\rho C} \right]^{-1}, & \text{otherwise,} \end{cases} \tag{S7}$$

$$T_C = \begin{cases} 0, & C = 0 \text{ or } \rho = 1 \\ \kappa_C \left[ \tau + \kappa_C \frac{1-\eta_C}{(1-\rho) C} \right]^{-1}, & \text{otherwise.} \end{cases} \tag{S8}$$

The above quantities appear as the testing expressions in $R_c$ given by Eq. (1) in the Methods section of the main text. It can be seen that $T_N$ and $T_C$ approach $\kappa_N/\tau$ and $\kappa_C/\tau$, respectively, as $C$ approaches $\infty$ and that both quantities approach 0 as $C$ approaches 0. Using these expressions, the derivation of $R_c$ and $R_0$ are identical to the derivation in Demers et al. (2023)[20]. See this reference for the steps leading to Eqs. (1) and (7) in the Methods section of the main text.
